# PILocarpine 1.25% Efficacy for management of Uncorrected Presbyopia (PILE-UP Study)

**DOI:** 10.1101/2024.10.04.24314877

**Authors:** Rohit Saxena, Vinay Gupta, T Velpandian, Tanya Nidhi, Himani Thakur, Jeewan S Titiyal, Namrata Sharma

## Abstract

**Purpose:** This study aimed to evaluate the efficacy and safety of pilocarpine 1.25% eye drops for improving near vision acuity in presbyopic individuals.

**Design:** Non-randomized prospective interventional trial

**Methods:** **This single-arm prospective interventional trial includes** fifty presbyopic individuals aged between 40 to 55, administrated pilocarpine 1.25% eye-drops daily once (9 am, Hour 0) for one month. Visual parameters, including distance corrected near visual acuity (DCNVA), near add power, amplitude of accommodation (AoA), and depth of focus (DoF) along with ocular biometric parameters were assessed at baseline and after one month at hour 3 (12 noon) and hour 6 (3 p.m.). Adverse effects were monitored.

**Results:** After one month of pilocarpine treatment, a significant improvement was observed in DCNVA (0.64±0.2 to 0.26±0.11, *P:<0*.*001*) along with a decrease in near add power (1.43±0.43 D to 0.42±0.22 D, *P:<0*.*001*). The AoA increased (3.23±0.74D to 3.92±0.93D, *P:0*.*005*), and DoF widened (0.72±0.18D to 0.81±0.26D, *P:0*.*038*). No change in ocular biometry parameters was observed. The change in DCNVA showed strong positive correlations with change in near add (r: 0.84, *P:<0*.*001*) and AoA (r: 0.66, *P:0*.*04*). Adverse effects were mild and did not lead to discontinuation.

**Conclusion:** Pilocarpine 1.25% eye drops demonstrated statistically significant improvements in DCNVA, near add, AoA, and DoF after one month of treatment. Pilocarpine 1.25% eye drops shown increase in the amplitude of accommodation and depth of focus; without any significant change in ocular biometry parameters. The use of pilocarpine 1.25% eye drops can be an alternative intervention for enhancing near vision acuity of presbyopic subjects.

## Introduction

Presbyopia is an age-related condition resulting in loss of the eye’s ability to focus on near objects due to progressive loss of accommodation with increasing age. Accommodation loss is likely because of loss of contraction power in ciliary body and viscoelasticity of the crystalline lens resulting in hardening of lens.^1^ Uncorrected presbyopia globally affect 1.8 billion people approximately, 826 million of whom had near vision impairment because of no, or inadequate, vision correction.^2^ It decrease the quality of life by affecting individual’s emotional well being and ability to do daily activities.^3^

The current options for correction of presbyopia include spectacle for near work e.g. single vision reading glasses, bifocal, multifocal progressive glasses; multifocal contact lens and some surgical techniques.^4-6^ But none of these methods able to restore dynamic range of accommodation.^7^ Multifocal glasses are often considers to be inconvenient by many and require need to direct the visual axes in a particular direction for adequate near vision.^8^

Pharmacological treatment for presbyopia has been under investigation recently.^9-12^ Pilocarpine is a cholinergic muscarinic receptor agonist, and may expand the diversity of existing strategies for treating presbyopia. It acts by enhancing depth of focus as well as accommodation for the treatment of presbyopia.^7,10^ Pilocarpine contracts ciliary muscles and iris sphincter muscles by binding to and activating muscarinic M3 receptors.^13^ Pupillary constriction caused by contraction of the iris sphincter create a pinhole effect that improves the ability to focus on near objects by increasing depth of focus.^14^ Also, accommodation is enhanced by contraction of ciliary muscles which also improve the near vision.

Pilocarpine 1.25% has been shown as a potential alternative for presbyopia treatment in recent studies,^10-12,15^ and has been approved by FDA^16^ as treatment of presbyopia. However current available studies are mainly in Caucasian population, its effect on the eyes with pigmented iris has not been evaluated. Pilocarpine demonstrates greater bioavailability in pigmented eyes, suggesting a biophase preference. Subjects with blue and brown eyes exhibited comparable levels of miosis when administered pilocarpine, yet the onset of peak effects was notably prolonged in the latter group.^17^ Furthermore, there exists a positive correlation between the density of pigmentation and the uptake capacity of pilocarpine.^18^ Major unaddressed questions related to pilocarpine for management of presbyopia are safety, optimum duration, and effect on ocular biometry parameters. There is lack of information on the efficacy and safety of pilocarpine 1.25% for treatment of presbyopia among pigmented eyes.

This study aims to evaluate the effect of pilocarpine 1.25% eye drops on near vision acuity and accommodation (efficacy outcomes); and associated side effect (if any) of pilocarpine 1.25% eye drops as measure by pupil size, ocular biometry, and occurrence of adverse events (safety outcomes).

### Methodology

The study was a prospective interventional trial [CTRI: REF/2022/07/056632] following the principles outlined in the Declaration of Helsinki. The study protocol received approval from the Institute Ethics Committee [Ref. No.: IEC-623/15.07.2022], and all participants provided written informed consent.

The inclusion criteria encompassed individuals aged 40 to 55 years, in good general health, and exhibiting both objective and subjective evidence of presbyopia. A thorough assessment of ocular and general health was conducted, including vision evaluation, slit-lamp examination for anterior segment health, cycloplegic refraction using tropicamide 1%, fundus examination and measurement of intraocular pressure. Emmetropes were defined as individuals with a spherical equivalent refractive error between -0.50 D and +0.50 D for distance in each eye, along with <0.75 D of astigmatism. Key inclusion criteria required participants to have high contrast uncorrected logMAR distance visual acuity better than or equal to 0.1, best corrected near visual acuity of N6 at 33cm in both eyes, intraocular pressure (IOP) between 10 to 21 mm of Hg, mesopic pupil size < 8.00 mm, and photopic pupil size > 3.00 mm. On the other hand, key exclusion criteria involved the presence of severe dry eye disease, corneal abnormalities, cataract, or any other retinal/ocular pathology, a history of ocular surgery and/or phakic intraocular lens implantation, migraine headaches, angle-closure glaucoma, abnormal pupil shape, anisocoria >1 mm, amblyopia, and known allergy to pilocarpine eye drops.

Throughout the study, participants received pilocarpine 1.25% eye drops once daily at 9 am in both eyes for one month, prepared by our in-house pharmacy compliant with good laboratory practice. Isotonic pilocarpine 1.25% eye drops (1.25 % of pilocarpine nitrate preserved with chlorbutol) were compounded from sterile dispensing facility of our pharmacy. Study visits were scheduled at baseline and a one-month follow-up. The data was recorded at 12 noon (Hour 3) and 3 pm (Hour 6). During the visits, objective and subjective refraction, along with near add, were performed. The patients’ acceptance of distance and near vision was recorded. Parameters noted included spherical equivalent refractive error, near correction, uncorrected visual acuity for both distance and near, best corrected visual acuity for both distance and near, and distance corrected near visual acuity. The near visual acuity was measured using near logMAR chart [ETDRS 2000 series (logarithmic chart calibrated for 33 cm), Precision Vision, Woodstock, IL, USA] at 33 cm in mesopic light conditions (10 lux at target). Additionally, optical biometry using partial interferometry optical biometer (IOL Master® 700, Zeiss, Germany) was conducted to measure axial length (AL), keratometry (Km), anterior chamber depth (ACD), vitreous chamber depth (VCD), and lens thickness (LT). The near point of accommodation was calculated using the RAF ruler, and the amplitude of accommodation (AoA) was determined as the inverse of this value. Pupil size was measured using automated, hand □ held, infrared pupillometer (NeurOptics PLR □ 200, Irvine, USA); and I-tracey (Tracey™ Technologies, Houston, TX) was employed to document depth of focus (DoF). Three readings were taken and median was recorded for purpose of analysis. The participants were asked to report any adverse effect such as headache, flashes of light etc. The individual was inquired about their response to the treatment, specifically regarding any challenges they experienced in performing routine near tasks and during close work or delicate tasks. This included questions about whether they encountered minimal or no difficulties in performing these tasks, as well as whether they were unable to carry out routine or delicate tasks even with the use of pilocarpine 1.25%. An anterior segment examination using slit lamp and central as well as peripheral retinal examination using indirect ophthalmoscopy was done at 1 month follow up.

After conducting a pilot study at the study centre from September 2022 to October 2022, we performed a conservative estimation of the effect size. To detect a minimum difference of 5 lines (0.5 logMAR visual acuity) on the logMAR near visual acuity chart, with a significance level of 0.05 and a power of 90%, sample size of 43 subjects is required. Accounting for an attrition rate of 15%, we determined that a total sample size of 50 subjects would be necessary.

Statistical analysis was conducted using IBM SPSS Statistics software, Version 25.0 (IBM Corp., Armonk, NY). The variables were represented in terms of mean and standard deviation. For a comprehensive examination of the data, linear mixed effects models were employed to address missing data at random and account for the correlation between the two eyes within each participant. Clustered data from both eyes of the same patient were combined using robust standard error.^19^ To assess the clinical significance of changes in variable characteristics, the P value was calculated using paired sample t-test for intra-group comparisons. P value of less than 0.05 was considered statistically significant.

## Results

The study enrolled 50 subjects with a mean age of 45.3 ± 5.1 years, of whom 45% were males. All the subjects had dark brown iris. There were no statistically significant differences observed in the baseline characteristics between the measurements taken at 12 pm and 3 pm at baseline (*P > 0*.*05* for all parameters). *(Table 1)*

**Table 1:** Mean value of baseline parameters of study subjects.

| Baseline Study Parameters | 12 noon | 3 pm |
| --- | --- | --- |
| UCVA (logMAR) | 0.04 ± 0.02 | 0.04 ± 0.02 |
| BCVA (logMAR) | 0.00 ± 0.00 | 0.00 ± 0.00 |
| DCNVA (logMAR) | 0.74 ± 0.2 | 0.76 ± 0.22 |
| BCNVA spectacles (logMAR) | 0.12 ± 0.01 | 0.12 ± 0.01 |
| SE Ref. error (distance) (D) | 0.08 ± 0.41 | 0.1 ± 0.32 |
| Near Addition (D) | 1.43 ± 0.43 | 1.46 ± 0.42 |
| IOP (mm of Hg) | 16.2 ± 2.5 | 15.9 ± 2.7 |
| Pupil size Mesopic (mm) | 5.3 ± 1.1 | 5.5 ± 1.3 |
| Pupil size photopic (mm) | 3.7 ± 0.6 | 3.8 ± 0.6 |
| Amplitude of accommodation (D) | 3.23 ± 0.74 | 3.14 ± 0.62 |
| Depth of focus (D) | 0.72 ± 0.18 | 0.68 ± 0.2 |
| Axial length (mm) | 23.59 ± 2.22 | 23.6 ± 2.1 |
| Mean Km (D) | 43.78 ± 3.7 | 43.85 ± 3.4 |
| ACD (mm) | 3.25 ± 0.33 | 3.26 ± 0.31 |
| LT (mm) | 3.41 ± 0.64 | 3.42 ± 0.7 |

UCVA: Uncorrected distance visual acuity; BCVA: best corrected distance visual acuity; DCNVA: distance corrected near visual acuity; SE: Spherical equivalent; IOP: Intraocular pressure; ACD: Anterior chamber depth; LT: lens thickness.

The mean follow-up time was 29.8 ± 1.6 days. At the one-month follow-up (12 noon), there was a statistically significant improvement in the logMAR distance corrected near visual acuity (DCNVA), which changed from 0.74 ± 0.2 at baseline to 0.36 ± 0.11 at one month *(P < 0*.*001)*. This improvement was accompanied by a decrease in the near add power, from 1.43 ± 0.43 D at baseline to 0.42 ± 0.22 D at one month *(P < 0*.*001)*. Additionally, an increase in the Amplitude of Accommodation was observed, changing from 3.23 ± 0.74 D to 3.92 ± 0.93 D *(P = 0*.*005)*, along with an increase in the Depth of Focus from 0.72 ± 0.18 D to 0.81 ± 0.26 D *(P = 0*.*038)*. No significant change observed in any other studied parameters. *(Table 2)*

**Table 2:** Mean characteristics of study subject at baseline and 1 month follow-up.

| Study parameters | Baseline (12 noon) | 1 month (12 noon) | P value |
| --- | --- | --- | --- |
| UCVA (logMAR) | 0.04 ± 0.02 | 0.06 ± 0.02 | 0.15 |
| BCVA (logMAR) | 0.00 ± 0.00 | 0.00 ± 0.00 | 1 |
| DCNVA (logMAR) | 0.74 ± 0.2 | 0.36 ± 0.11 | <0.001 |
| BCNVA spectacles (logMAR) | 0.12 ± 0.01 | 0.11 ± 0.02 | 0.25 |
| SE Ref. error (distance) (D) | 0.08 ± 0.41 | 0.13 ± 0.39 | 0.12 |
| Near Addition (D) | 1.43 ± 0.43 | 0.42 ± 0.22 | **<0.001** |
| IOP (mm of Hg) | 16.2 ± 2.5 | 16.8 ± 3.5 | 0.2 |
| Pupil size Mesopic (mm) | 5.3 ± 1.1 | 5.13 ± 1.4 | 0.066 |
| Pupil size photopic (mm) | 3.7 ± 0.6 | 3.51 ± 0.5 | 0.052 |
| Amplitude of accommodation (D) | 3.23 ± 0.74 | 3.92 ± 0.93 | **0.005** |
| Depth of focus (D) | 0.72 ± 0.18 | 0.81 ± 0.26 | **0.038** |
| Axial length (mm) | 23.59 ± 2.22 | 23.62 ± 2.24 | 0.11 |
| Mean Km (D) | 43.78 ± 3.7 | 43.89 ± 4.1 | 0.14 |
| ACD (mm) | 3.25 ± 0.33 | 3.27 ± 0.39 | 0.08 |
| LT (mm) | 3.41 ± 0.64 | 3.43 ± 0.81 | 0.071 |

UCVA: Uncorrected distance visual acuity; BCVA: best corrected distance visual acuity; DCNVA: distance corrected near visual acuity; SE: Spherical equivalent; IOP: Intraocular pressure; ACD: Anterior chamber depth; LT: lens thickness.

Furthermore, there was a strong positive correlation between the change in DCNVA & the change in Near Add *(r: 0*.*84, P < 0*.*001);* and change in DCNVA & change in amplitude of accommodation (AoA) *(r: 0*.*66, P: 0*.*04)*. No strong corelation observed between change in DCNVA & change in depth of focus *(r: 0*.*32. P: 0*.*075*); and change in near add & change in depth of focus *(r: 0*.*28, P: 0*.*062)*. However, a strong corelation found between change in AoA & change in near add *(r: 0*.*61, P: 0*.*048)*; and change in AoA & change in depth of focus *(r: 0*.*71, P: 0*.*036)*.

At 1 month follow-up, significant difference was observed in DCNVA *(P: .046)*, amount of near add *(P: .031)*, and amplitude of accommodation *(P: .048)*, between hour 3 (12 noon) and hour 6 (3 pm) post instillation of pilocarpine 1.25% drop. *(Table 3)* However all these 3 parameters remained significantly better at 1 month compared to the baseline values at 6 hour (3 pm) *(P < 0*.*001). (Table 4)*

**Table 3:** Mean characteristics of study subjects at hour 3 and hour 6.

| **1 month follow-up** | **12 noon (Hour 3)** | **3 pm (Hour 6)** | **P value** |
| --- | --- | --- | --- |
| UCVA (logMAR) | 0.06 ± 0.02 | 0.04 ± 0.04 | 0.25 |
| BCVA (logMAR) | 0.00 ± 0.00 | 0.00 ± 0.00 | 1 |
| DCNVA (logMAR) | 0.36 ± 0.11 | 0.44 ± 0.14 | **0.046** |
| BCNVA spectacles (logMAR) | 0.11 ± 0.02 | 0.12 ± 0.02 | 0.17 |
| SE Ref. error (distance) (D) | 0.13 ± 0.39 | 0.08 ± 0.4 | 0.08 |
| Near Addition (D) | 0.42 ± 0.22 | 0.52 ± 0.26 | **0.031** |
| IOP (mm of Hg) | 16.8 ± 3.5 | 16.6 ± 4.1 | 0.22 |
| Pupil size Mesopic (mm) | 5.13 ± 1.4 | 5.39 ± 1.1 | 0.073 |
| Pupil size photopic (mm) | 3.51 ± 0.5 | 3.72 ± 0.8 | 0.01 |
| Amplitude of accommodation (D) | 3.92 ± 0.93 | 3.64 ± 0.62 | **0.048** |
| Depth of focus (D) | 0.81 ± 0.26 | 0.76 ± 0.21 | 0.1 |
| Axial length (mm) | 23.62 ± 2.24 | 23.61 ± 2.7 | 0.36 |
| Mean Km (D) | 43.89 ± 4.1 | 43.75 ± 3.9 | 0.14 |
| ACD (mm) | 3.27 ± 0.39 | 3.25 ± 0.44 | 0.13 |
| LT (mm) | 3.43 ± 0.81 | 3.43 ± 0.74 | 0.16 |

UCVA: Uncorrected distance visual acuity; BCVA: best corrected distance visual acuity; DCNVA: distance corrected near visual acuity; SE: Spherical equivalent; IOP: Intraocular pressure; ACD: Anterior chamber depth; LT: lens thickness.

**Table 4:** Mean characteristics of study subjects at baseline (3pm) versus 1 month follow-up (3pm)

| **1 month follow-up** | **Baseline (3 pm)** | **1 month follow-up**<br>**3 pm (Hour 6)** | **P value** |
| --- | --- | --- | --- |
| UCVA (logMAR) | 0.04 ± 0.02 | 0.04 ± 0.04 | 0.72 |
| BCVA (logMAR) | 0.00 ± 0.00 | 0.00 ± 0.00 | 1 |
| DCNVA (logMAR) | 0.76 ± 0.22 | 0.44 ± 0.14 | **>0.001** |
| BCNVA spectacles (logMAR) | 0.12 ± 0.01 | 0.12 ± 0.02 | 0.54 |
| SE Ref. error (distance) (D) | 0.1 ± 0.32 | 0.08 ± 0.4 | 0.17 |
| Near Addition (D) | 1.46 ± 0.42 | 0.52 ± 0.26 | **>0.001** |
| IOP (mm of Hg) | 15.9 ± 2.7 | 16.6 ± 4.1 | 0.15 |
| Pupil size Mesopic (mm) | 5.5 ± 1.3 | 5.39 ± 1.1 | 0.055 |
| Pupil size photopic (mm) | 3.8 ± 0.6 | 3.72 ± 0.8 | 0.062 |
| Amplitude of accommodation (D) | 3.14 ± 0.62 | 3.64 ± 0.62 | **>0.001** |
| Depth of focus (D) | $0.68 \pm 0.2$ | $0.76 \pm 0.21$ | *0.048* |
| Axial length (mm) | $23.6 \pm 2.1$ | $23.61 \pm 2.7$ | *0.48* |
| Mean Km (D) | $43.85 \pm 3.4$ | $43.75 \pm 3.9$ | *0.2* |
| ACD (mm) | $3.26 \pm 0.31$ | $3.25 \pm 0.44$ | *0.31* |
| LT (mm) | $3.42 \pm 0.7$ | $3.43 \pm 0.74$ | *0.12* |

UCVA: Uncorrected distance visual acuity; BCVA: best corrected distance visual acuity; DCNVA: distance corrected near visual acuity; SE: Spherical equivalent; IOP: Intraocular pressure; ACD: Anterior chamber depth; LT: lens thickness.

The patient satisfaction response to treatment summarized in *Table 5*.

**Table 5:** Patient satisfaction response to treatment.

| **Response** | **Number** |
| --- | --- |
| No difficulty in doing routine near work as well as fine work | 30/50 (60%) |
| Easily doing routine near work, but slight difficulty in doing fine work | 15/50 (30%) |
| Slight difficulty in doing routine near work, can't do fine work | 4/50 (8%) |
| Can't do routine near work properly | 1/50 (2%) |

The adverse effects reported by patient includes headache (8/50, 16%), eye pain (1/50, 2%), visual blur (2/50, 4%) and irritation (2/50, 4%). Symptoms are during initial days of treatment and was not severe. No patient discontinued using pilocarpine 1.25% due to adverse effect. No relevant anterior segment as well as retinal changes was observed at 1 month follow up when accessed through slit-lamp examination and dilated fundus examination respectively.

## Discussion

In this prospective interventional study, we assessed the impact of 1.25% pilocarpine eye drops on both near vision acuity and accommodation, while also monitoring any potential adverse events associated with the drug. After one month, an improvement in Distance Corrected Near Visual Acuity (DCNVA) at hour 3, along with a simultaneous reduction in the Near-Add required by the patient was observed. The term “near add” refers to the additional optical power needed to achieve clear near vision, typically in the form of reading glasses or bifocal/multifocal lenses. By contracting the ciliary muscle, pilocarpine increases the eye’s ability to adjust its focus for near vision tasks,^18^ reducing the need for additional optical correction (near add) to achieve clear near vision.

Our study showed a significant improvement in DCNVA, AoA, DoF along with reduction in near add requirement after 1 month of treatment as compared to baseline. GEMINI-1 Phase 3 Randomized Controlled Trial (RCT)^12^, pilocarpine 1.25% demonstrated superiority over the vehicle in terms of DCNVA on day 30, hours 3, and 6. Specifically, on day 30, hour 3, 30.7% of participants in the pilocarpine 1.25% group showed an improvement of 3 or more lines in DCNVA, compared to only 8.1% in the vehicle group. At hour 6, these percentages were 18.4% and 8.8%. In our study 44% of participant at hour 3 and 30% of participant at hour 6 showed an improvement of 0.5 logMAR (5 lines on logMAR scale). Our study revealed a significant decrease in logMAR DCNVA at hour 6 compared to hour 3 on day 30. This suggests that the effect of pilocarpine on near vision acuity may start to diminish over time after the initial application, and the efficacy for near addition tends to be higher during the first few hours following application; though the effect of pilocarpine 1.25% was retained at hour 6 when compared to baseline The VIRGO trial had demonstrated significant benefit with Pilocarpine 1.25% twice daily compared to placebo.^15^ It is possible that twice daily dose may provide more sustained effect.

There was no change observed after 1 month of using pilocarpine 1.25% in Best-Corrected Visual Acuity (BCVA) for distance. This finding aligns with the GEMINI-1 Phase 3 RCT, where no participants with a DCNVA improvement of 3 lines or more experienced a loss of more than 5 letters in BCVA for distance on day 30.^12^

In addition to evaluating near vision acuity, our study also compared the amplitude of accommodation and depth of focus at baseline and after 1 month of treatment. We observed a significant increase in the amplitude of accommodation and depth of focus at day 30, hour 3. The increase in the amplitude of accommodation after 1 month of using pilocarpine 1.25% is likely due to the drug’s pharmacological effects on the eye’s anatomy and physiology. These findings indicate that pilocarpine’s mechanism of action involves pupillary constriction, which leads to an increase in depth of focus, as well as contraction of the ciliary muscle, resulting in an increased amplitude of accommodation. When the ciliary muscle contracts, it causes the lens of the eye to change its shape, becoming more rounded. This increased curvature of the lens enhances its refractive power, enabling the eye to focus on nearby objects. Also, a greater depth of focus, such as that achieved through the use of pilocarpine eye drops, can enhance near vision by expanding the range of distances at which near objects can be seen clearly without straining the eyes or constantly adjusting the focus.

The strong correlation observed between the change in near add and the amplitude of accommodation suggests that the improvement in near vision acuity is directly related to the increased ability of the eye to accommodate for near objects. As pilocarpine induces pupillary constriction and ciliary muscle contraction, it enhances the eye’s accommodative response, resulting in improved near vision and a decreased reliance on additional optical correction (near add) for near tasks.

Despite independent improvements in both near visual acuity and depth of focus, the study did not find a statistical correlation between the two. In other words, the improvement in near visual acuity did not necessarily correlate with the increase in depth of focus. While pilocarpine eye drops can lead to improvements in near visual acuity and depth of focus independently, other factors or mechanisms may be responsible for these effects,^7,20^ and they might not be directly related to each other. The study’s findings provide valuable insights into the effects of pilocarpine on various visual parameters, but further research may be needed to fully understand the underlying mechanisms and potential interactions between near visual acuity and depth of focus in response to pilocarpine treatment.

Notably, the daily use of 1.25% pilocarpine was generally well-tolerated, with minimal side effects. Ninety percent of participants in our study indicated experiencing either no difficulties or only minimal challenges when engaging in both routine near work and at close-distances/more-intricate tasks with using pilocarpine 1.25%. In this study, despite the pupillary constriction induced by pilocarpine, there was no significant change in the size of the pupil measured in both dim and well-lit conditions at the end of the 1-month treatment period. This finding suggests that the effect of pilocarpine on pupil size was likely not sustained over the long term or was not significant enough to produce a measurable change in pupil size in the studied population. The lack of significant change in pupil size after 1 month of pilocarpine use may be considered beneficial, as excessive or prolonged pupillary constriction could potentially lead to visual disturbances and discomfort, especially in low-light conditions. While earlier research had documented instances of retinal detachment associated with the use of pilocarpine 1.25% for presbyopia,^21-22^ our study did not identify any occurrences of retinal detachment or other retinal pathologies. However, individual responses to medications should always be monitored by healthcare professionals to ensure safety and optimal treatment outcomes.

This study has certain limitations. Firstly, being a non-randomized trial, there is a potential for biases when compared to blinded randomized controlled trials. Secondly, the sample size is modest; a larger cohort studied over an extended period would offer deeper insights into the drug’s efficacy and safety. However, this study stands out as the pioneering research documenting the efficacy of pilocarpine in eyes with pigmented iris, setting a foundation for future studies. A key strength of our study is the quantification of ocular biometry parameters, amplitude of accommodation and depth of focus among participants administered with pilocarpine 1.25% for presbyopia. To our current understanding, this is the first study quantifying accommodation and depth of focus and correlating them with the adjustments in near add required and enhancements in near visual acuity; among individuals given pilocarpine 1.25% for presbyopia.

In conclusion, the study’s findings suggest that the use of pilocarpine 1.25% eye drops can be an alternative intervention for enhancing near vision acuity in the presbyopic subjects. Pilocarpine 1.25% eye drops shows statistically significant improvement in uncorrected near visual acuity; increase in the amplitude of accommodation and depth of focus among studied presbyopic individuals, without any significant change in ocular biometry parameters. No occurrence of retinal detachment or any other severe adverse effect was observed in our study. However further research and individual assessments would be necessary to fully understand and optimize the benefits of pilocarpine for near vision improvement in different clinical contexts.

*All authors have completed and submitted the ICMJE form for disclosure of potential conflicts of interest. Funding/Support: None. Financial disclosure: No financial disclosure to make for any author(s)*.

## Data Availability

Data will be made available on reasonable request.

